# Thoracic Aortic Disease in Genetic Syndromes Beyond Established Aortopathy Genes

**DOI:** 10.64898/2026.09.04.26362279

**Authors:** David R. Murdock, Bobbi McGivern, Dongchuan Guo, Kirsty McWalter, Dianna M. Milewicz

**Author notes:** Address for correspondence: Dianna M. Milewicz, MD, PhD UTHealth Houston, 6431 Fannin St, MSB 6.100, Houston, Texas 77030.

## Abstract

**Background:** Heritable thoracic aortic aneurysms and dissections (HTAD) are caused by rare variants in up to 42 genes, but many affected individuals and families remain genetically unsolved. We sought to determine if aortic phenotypes occurred in cases with *de novo* pathogenic variants in known syndromic genes.

**Methods:** We queried a clinical exome/genome sequencing database for individuals with *de novo* pathogenic or likely pathogenic variants (PVs) and aortic Human Phenotype Ontology terms. Genes with PVs present in three or more unrelated cases with aortic phenotypes were further evaluated using published evidence, genome-wide rare-variant burden analyses in the UK Biobank and All of Us, and case-level data from independent thoracic aortic disease (TAD) cohorts.

**Results:** Among 250 individuals with *de novo* PVs and aortic phenotypes, 32 (12.8%) had PVs in genes included on commercial HTAD panels and were excluded from further analyses. The remaining 218 individuals had PVs in 144 genes, including 16 genes identified in three or more unrelated individuals. Genes involved in chromatin regulation and RAS/mitogen-activated protein kinase (MAPK) signaling, as well as several genes associated with overgrowth syndromes, were recurrently observed. Across biobank analyses, independent TAD cohorts, and published reports, the strongest evidence for aortic involvement was observed for *DNMT3A, NF1, PTPN11, PPP1CB, NSD1, ANKRD11, ADNP,* and *ABL1*, including significant rare-variant burden associations for three of these genes. Aortic phenotypes also occurred across multiple Coffin-Siris/BAF-complex genes.

**Conclusions:** Aortic phenotypes occur across a diverse range of genetic syndromes beyond established HTAD genes. These findings support broader genomic testing in TAD, expansion of HTAD panels to include additional syndromic genes, and consideration of aortic surveillance in select syndromic disorders.

## Background

Thoracic aortic disease (TAD) is a major cause of premature morbidity and mortality, with acute aortic dissection often resulting in sudden death[1,2]. Many of these events are potentially preventable when individuals at increased genetic risk are identified before dissection occurs. A genetic diagnosis can prompt aortic imaging and surveillance, identify at-risk relatives through cascade testing, and enable elective repair of enlarging aneurysms. Marfan syndrome provides a model for this approach, as earlier recognition, cardiovascular surveillance, medical management, and prophylactic aortic root repair have substantially improved survival[3,4]. These advances underscore the clinical importance of recognizing the full spectrum of genetic disorders that predispose to aortic aneurysm and dissection.

Pathogenic variants in a curated group of 11 heritable thoracic aortic disease (HTAD) genes involved in extracellular matrix integrity (*FBN1, COL3A1, LOX*), smooth muscle cell contraction (*ACTA2, MYH11, MYLK, PRKG1*), and TGF-β signaling (*TGFBR1, TGFBR2, TGFB2, SMAD3*), along with many more putative genes that have yet to be curated, contribute to HTAD[5,6]. These discoveries have shaped clinical practice and led to targeted gene panels and structured aortic surveillance and clinical management recommendations for affected individuals and at-risk relatives[2].

Nevertheless, a substantial proportion of individuals with TAD remain unexplained, with pathogenic/likely variants (PVs) in the 11 validated genes accounting for only 30% of HTAD families and 10% of dissections [5,6]. Concurrently, the increasing use of clinical exome/genome sequencing (ES/GS) in individuals with neurodevelopmental, overgrowth, and multisystem disorders has expanded the recognized phenotypic spectrum of many Mendelian conditions.

Aortic dilation and aneurysm have been described in disorders not traditionally classified as primary aortopathies, including RAS/MAPK pathway disorders, such as Noonan syndrome, and chromatin-related syndromes, including Kabuki, Sotos, and Tatton-Brown-Rahman syndromes [7–10]. However, in many cases, these vascular findings have been reported in small series or individual case reports, and their broader contribution to unexplained aortic phenotypes remains uncertain.

Whether additional syndromic genes, particularly those identified through broad genomic sequencing of individuals with multisystemic phenotypes, contribute meaningfully to TAD has not been systematically evaluated. A phenotype-driven genomic approach offers an opportunity to address this gap by identifying individuals with documented thoracic aortic involvement who lack PVs in HTAD genes. In this study, we queried a large clinical genomic sequencing database to identify individuals with *de novo* PVs and documented TAD-specific Human Phenotype Ontology (HPO) terms, excluding those with variants in genes included in HTAD diagnostic panels [11]. We then characterized the implicated genes, including their established associations with Mendelian diseases and evidence linking them to TAD. Through this systematic evaluation, we aimed to define the spectrum of TAD phenotypes associated with established genetic syndromes and thus inform future approaches to genetic testing and aortic surveillance.

## Methods

### Primary GeneDx Cohort

We conducted a retrospective cohort study of individuals with aortic phenotypes identified through clinical diagnostic ES/GS at GeneDx. The initial phenotype-query cohort included 1,216 individuals with at least one *de novo* variant and at least one aortic or arterial HPO term (Table S1) with testing reported between March 2013 and October 2023. HPO terms were assigned to each individual from the abstraction of the medical records and test requisition provided by the ordering clinician at the time of genetic testing, using Neji concept recognition, followed by manual review by a laboratory clinician [12]. To describe the clinical characteristics of the cohort, we mapped clinical phenotypes to curated HPO terms using a standardized internal GeneDx framework. We grouped individual HPO terms according to the HPO hierarchical structure (https://hpo.jax.org/), assigning each term to its corresponding higher-level parent phenotype category. This allowed the aggregation of related phenotypic features into broader clinical phenotype groups, whose prevalence could then be summarized (Table S2). Eligible individuals for the primary analytic cohort met all the following criteria: 1) presence of a *de novo* PV identified and reported at the time of clinical ES/GS, 2) an aortic phenotype documented by the ordering clinician and included in a prespecified list of abstracted HPO terms, and 3) absence of a PV in any of 42 unique genes represented across three major diagnostic laboratory HTAD panels (Table S3).

### Genomic Sequencing and Variant Interpretation

For the ES cases, using genomic DNA from the proband and parent(s), the exonic regions and flanking splice junctions of the genome were captured using the SureSelect Human All Exon V4 (50 Mb), the Clinical Research Exome kit (Agilent Technologies, Santa Clara, CA), or the IDT xGen Exome Research Panel v1.0 (Integrated DNA Technologies, Coralville, IA). For the GS cases, PCR-free whole genome sequencing libraries were prepared using the Illumina DNA PCR-Free Library prep kit (San Diego, CA). In all cases, massively parallel (NextGen) sequencing was performed on an Illumina system with 150 bp paired-end reads. Reads were aligned to the human genome build GRCh37/UCSC hg19 and analyzed for sequence variants using a custom-developed analysis tool. In some cases, only genes of interest from ES, as selected by the ordering clinician, were included in the analysis. Reported variants were confirmed, if necessary, by an appropriate orthogonal method in the proband and in selected relatives. For both ES and GS cases, additional sequencing technology and a variant interpretation protocol have been previously described [13]. Only variants classified as pathogenic or likely pathogenic at the time of reporting according to American College of Medical Genetics and Genomics (ACMG) criteria were included[14]. The general assertion criteria for variant classification are publicly available on the GeneDx ClinVar submission page (http://www.ncbi.nlm.nih.gov/clinvar/submitters/26957/). *De novo* status was confirmed by parental testing when available. Genetically inferred ancestry was assigned by projecting individual genotypes onto the 1000 Genomes Project reference populations (www.internationalgenome.org) using principal component analysis, followed by statistical clustering.

### Gene Curation

Genes were aggregated across all qualifying probands in the GeneDx cohort, and recurrence was defined as the presence of *de novo* PVs in three or more unrelated individuals. Mendelian disease associations were defined using the Online Mendelian Inheritance in Man (OMIM) database[15]. Published associations with TAD were identified through targeted literature review using PubMed.

### Biobank Analyses

To evaluate independent evidence for genes recurrently identified in the primary GeneDx cohort, rare-variant burden analyses were performed using GS data from the UK Biobank (UKB) and the All of Us Research Program (AoU), comprising 4,889 TAD cases and 757,960 controls (Table S5)[16,17]. Aortic disease phenotypes were defined using diagnosis and procedure codes as detailed in the Supplemental Methods and Table S6. Controls excluded individuals with aortic disease or congenital/chromosomal abnormalities. Rare predicted damaging missense variants were defined as having gnomAD minor allele frequency (MAF) <8.16 × 10, REVEL >0.649, and AlphaMissense >0.2543 [18]. These thresholds were derived from a biobank-based analysis to identify rare missense variants associated with increased risk of thoracic aortic dissection and subsequently validated in additional cohorts. Predicted loss-of-function (LOF) variants were restricted to the same MAF threshold and included stop-gain, canonical splice-site, and frameshift variants in genes expected to be LOF-intolerant. Gene-based burden testing of missense, LOF, and combined missense and LOF variants was performed separately in the UKB and AoU using Firth-penalized logistic regression, adjusted for age, sex, and genetic principal components [19]. Cohort-specific results were combined using a fixed-effects inverse-variance weighted meta-analysis [20]. Benjamini-Hochberg false discovery rate (FDR) correction was applied to all available biobank burden analyses before selecting the most significant result for each gene [21]. Additional variant filtering, quality control, and analytical details are provided in the Supplemental Methods.

### Independent Case-Level Evidence

ES/GS was performed on DNA samples from unsolved affected individuals in additional UTHealth cohorts, including affected probands from 485 unrelated HTAD families, 564 individuals from the Early-Onset Sporadic Thoracic Aortic Dissection (ESTAD) cohort, and probands with thoracic aortic disease from 67 trios. These cohorts were queried for rare, potentially deleterious variants in genes recurrently identified in the primary GeneDx cohort.

Qualifying variants included missense variants with a minor allele frequency <5 × 10 in gnomAD v4.1.1 and REVEL >0.5, as well as in-frame insertions and deletions and predicted LOF variants meeting the same allele-frequency threshold. Additional sequencing, variant- filtering, and cohort details, including the rationale for these thresholds, are provided in the Supplemental Methods.

## Results

### Cohort Characteristics

The initial GeneDx phenotype-query cohort included 1,216 individuals with at least one *de novo* variant and at least one aortic or arterial HPO term (Table S1). Overall, these individuals had multisystem clinical findings, with 89.8% having an abnormality of the musculoskeletal system, 57.7% neurodevelopmental delay, 54.3% multiple congenital anomalies, and 32.1% with congenital heart disease (Table S2). Among the 250 individuals with one or more *de novo* PVs, 32 had one or more PVs in a gene included in the 42-gene union of commercial HTAD panels (Table S4). Thus, HTAD-panel genes accounted for 12.8% of *de novo* PV-positive individuals in this selected cohort, whereas the majority had PVs in genes outside the current commercial HTAD panel content. After excluding these cases, the final analytic cohort comprised 218 individuals with documented aortic phenotypes and reported *de novo* PVs in non-HTAD panel genes (two individuals had PVs in two distinct genes; Figure 1). The cohort was predominantly pediatric (83.9%), male (67.4%), and of European genetically inferred ancestry (49.1%).

**Figure 1.**
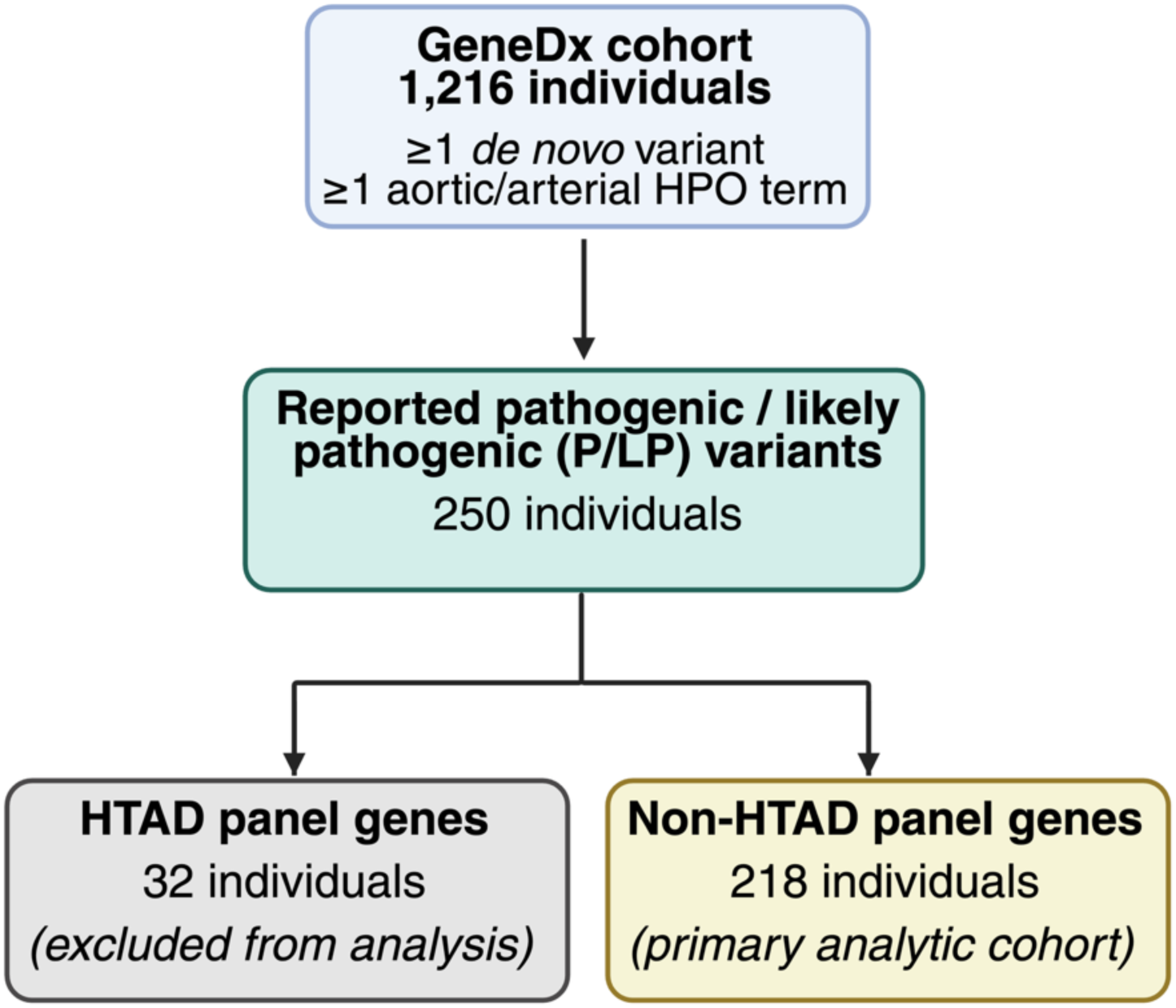
Study design and cohort assembly. Flow diagram summarizing selection of the final analytic cohort, including restriction to individuals with *de novo* pathogenic or likely pathogenic variants (PVs) and exclusion of PVs in genes represented on commercial HTAD panels.

Thoracic aortic aneurysm was the most common vascular phenotype (78.9%), with aortic root aneurysm most prevalent within that category (49.5%), and arterial dissection (4.1%), arterial tortuosity (2.8%), and aortic dissection (0.5%) were observed less frequently (Table 1, Table S7). Individuals in the final analytic cohort had multisystem disorders with an average of 28.7 clinical findings (i.e., HPO terms) per individual (range from 3 to 107) with the most common extra-vascular phenotypes reported as: global developmental delay (60.1%), generalized hypotonia (40.4%), delayed speech and language development (32.1%), abnormal facial shape (30.3%), and gastroesophageal reflux (25.7%) (Table S7).

**Table 1.** Demographic and vascular phenotypic characteristics of individuals with *de novo* PVs in non-HTAD panel genes as reported by clinical ES/GS.

| Characteristic | N (%) |
| --- | --- |
| Total individuals | 218 (100) |
| Unique genes | 144 |
| **Sex** |  |
| Male | 147 (67.4%) |
| Female | 71 (32.6%) |
| **Age at Testing (Years)** |  |
| 0–17 | 183 (83.9%) |
| 18–60 | 35 (16.1%) |
| **Genetically Inferred Ancestry** |  |
| European | 107 (49.1%) |
| Admixed American / Latino | 33 (15.1%) |
| African | 19 (8.7%) |
| Middle Eastern | 11 (5.0%) |
| East Asian | 5 (2.3%) |
| South Asian | 3 (1.4%) |
| Not analyzed | 40 (18.3%) |
| **Vascular Phenotype<sup>a</sup>** |  |
| **Thoracic aortic aneurysm (TAA) phenotype total (78.9%)** |  |
| Aortic root aneurysm (HP:0002616) | 108 (49.5%) |
| Ascending aortic aneurysm (HP:0004970) | 60 (27.5%) |
| Aortic arch aneurysm (HP:0005113) | 2 (0.9%) |
| Thoracic aortic aneurysm (not otherwise specified) (HP:0012727) | 2 (0.9%) |
| Descending thoracic aorta aneurysm (HP:0004959) | 0 (0%) |
| **Aortic aneurysm (not otherwise specified) (HP:0004942)** | **45 (20.6%)** |
| **Aortic/Arterial dissection phenotype total (4.1%)** |  |
| Arterial dissection (HP:0005294) | 4 (1.8%) |
| Aortic dissection (HP:0002647) | 3 (1.4%) |
| Ascending aortic dissection (HP:0004933) | 1 (0.5%) |
| Coronary artery dissection (HP:0006702) | 1 (0.5%) |
| Descending aortic dissection (HP:0012499) | 0 (0%) |
| Carotid artery dissection (HP:0012158) | 0 (0%) |
| **Aortic tortuosity (HP:0006687)** | **6 (2.8%)** |
| **Aortic rupture (HP:0031649)** | **1 (0.5%)** |

<sup>a</sup>Categories are not mutually exclusive.

### Gene Distribution and Recurrence

*De novo* PVs were distributed across 144 genes, demonstrating substantial genetic heterogeneity within the cohort (Tables S8). Most genes were observed in only a single individual, whereas a smaller subset recurred in multiple unrelated individuals. Overall, 106 genes (73.6%) were identified in one individual, 22 genes (15.2%) were identified in two individuals, and 16 genes (11.1%) were identified in three or more unrelated individuals.

The most frequently observed recurrent genes were *NSD1* and *PTPN11* (n=8 each), followed by *ANKRD11* (n=7), *ADNP* and *DNMT3A* (n=5 each), and *ABL1*, *ARID1B*, *CREBBP*, and *NF1* (n=4 each). Seven additional genes, *DPF2*, *KAT6A*, *PACS1*, *PIK3CA*, *PPP1CB*, *SRCAP*, and *ZNF292*, were identified in three individuals each (Figure 2). Variant-level details for the *de novo* PVs identified in the GeneDx cohort are provided in Table S9. Two individuals each carried two PVs: one with *CREBBP* and *ROBO2* variants, and the other with *ADNP* and *KAT6A* variants. Recurrent genes were distributed across several functional categories, including chromatin regulation, RAS/MAPK signaling, and syndromic neurodevelopmental or overgrowth disorders.

**Figure 2.**
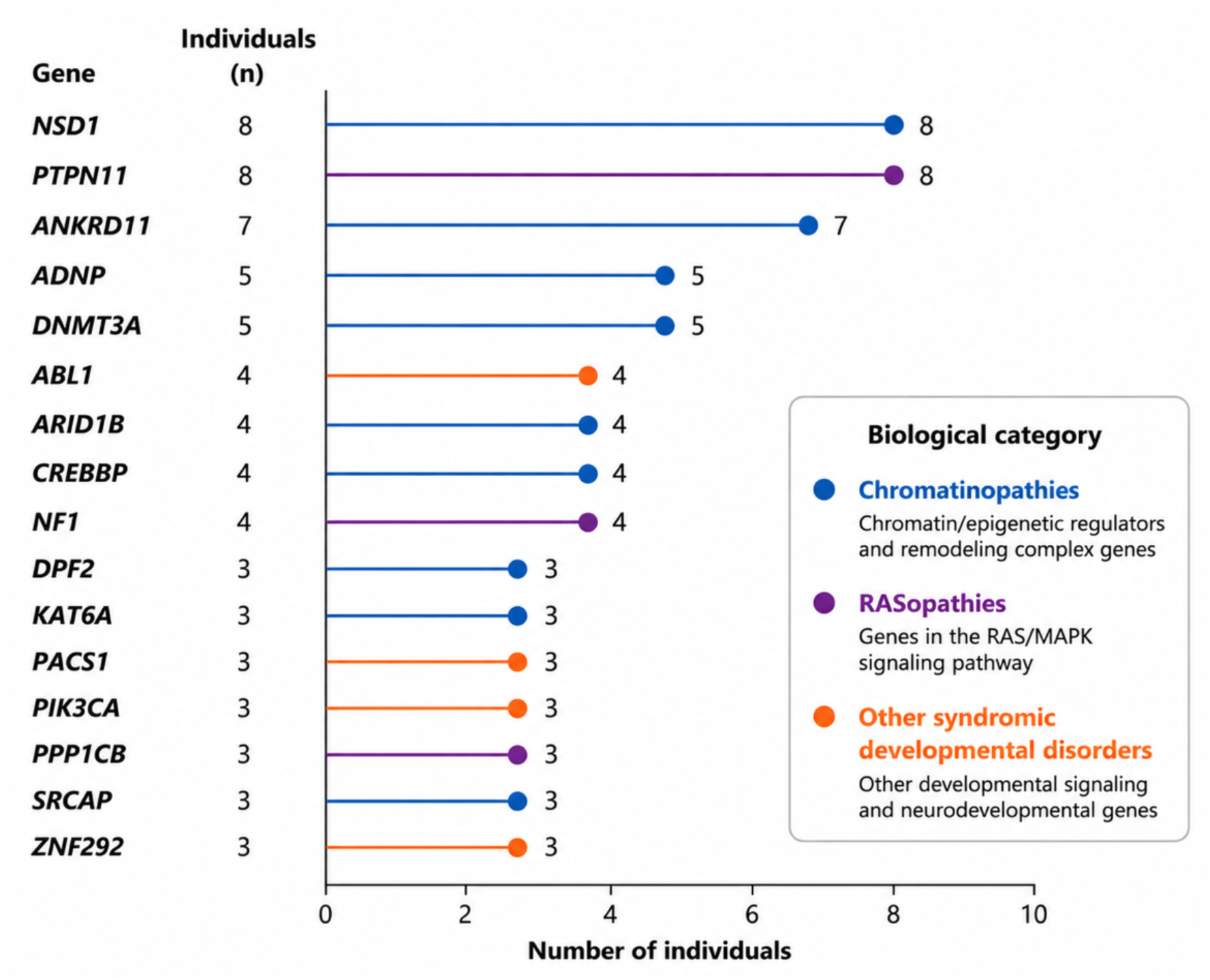
Syndromic genes with pathogenic variants associated with aortic phenotypes. Genes identified in three or more unrelated individuals with confirmed *de novo* PVs are ranked by recurrence and grouped into three major biological categories: chromatinopathies, RASopathies, and other syndromic developmental disorders.

Notably, several recurrent *de novo* PVs were observed in the cohort. The *PACS1* (NM_018026.3) c.607C>T (p.Arg203Trp) variant was identified in three unrelated individuals. Two of the three individuals with *PPP1CB* (NM_206876.1) PVs harbored the recurrent c.146C>G (p.Pro49Arg) variant, which has also been reported in a 2-year-old child with a dilated aortic root[22]. Finally, two individuals shared the *WT1* (NM_024426.4) c.1301G>A (p.Arg434His) variant, alternatively reported as c.1097G>A (p.Arg366His) depending on the transcript used.

### Biobank and Case-Level Evidence for Novel Syndromic Genes Associated with Aortic Phenotypes

For the 16 genes with three or more *de novo* cases with aortic phenotypes in the primary GeneDx cohort, we sought additional evidence for an association with TAD in broader, primarily adult cohorts (Table S5). We first evaluated rare-variant burden in TAD cases versus controls using UKB and AoU. After multiple-testing correction across UKB, AoU, and meta-analysis tests, three associations remained significant at an FDR <0.05 (Table S10). The strongest biobank burden evidence was observed for *NF1*, with LOF variants enriched among individuals with aortic dissection in the combined UKB/AoU meta-analysis (OR, 9.36; 95% CI, 2.74–32.03; *P* = 3.63×10 ; FDR-adjusted *P* = 0.021). One additional *NF1* variant was identified in an HTAD family. *PPP1CB* missense variants were enriched among individuals with thoracic aortic aneurysms in UKB (OR, 193.83; 95% CI, 15.46–1461.41; *P* = 6.62×10 ; FDR-adjusted *P* = 0.021). *ADNP* LOF variants were enriched among individuals with thoracic aortic aneurysms in AoU (OR, 25.54; 95% CI, 4.81–91.42; *P* = 0.001; FDR-adjusted *P* = 0.021).

We next examined independent, clinically ascertained TAD cohorts for additional case-level evidence. Qualifying variants were required to have a minor allele frequency <5 × 10 in gnomAD v4.1.1 and included predicted LOF variants, in-frame insertions or deletions, and missense variants with a REVEL score >0.5. Multiple additional cases were identified for *PTPN11* (*n* = 5), *KAT6A* (*n* = 5), *CREBBP* (n = 4), *DNMT3A* (*n* = 4), *PIK3CA* (*n* = 3), and *SRCAP* (*n* = 2) across our ESTAD and HTAD cohorts (Table S11). Notably, the *DNMT3A* (NM_022552.5) p.Arg635Gln variant identified in a sporadic aortic dissection case was the same variant previously reported in an individual with Tatton-Brown-Rahman syndrome and aortic root dilation[10]. Additional case-level observations were identified for *NF1*, *ABL1*, *DPF2*, *PACS1*, and *ZNF292*, each represented by a single individual in the additional cohorts. The integrated evidence for the 16 recurrent genes, including GeneDx recurrence, biobank burden results, external case-level observations, and published evidence of aortic disease, is summarized in Table S10.

## Discussion

Genes with rare variants that increase the risk for TAD have traditionally been identified using large HTAD families with multiple affected members due to the extensive genetic heterogeneity for the disease. Despite the success of this approach, PVs in established HTAD genes account for only a subset of individuals with TAD [6]. Using this unique cohort to assess *de novo* variants in cases with aortic phenotypes, we identified substantial genetic heterogeneity among cases, even after excluding genes represented on commercial HTAD panels. The final analytic cohort included 218 individuals with *de novo* PVs distributed across 144 non-panel genes, with 16 genes recurrently observed in three or more unrelated individuals. Among the 253 *de novo* PV-positive individuals in the parent cohort, only 32 (12.8%) had PVs in one of the 42 genes included in one or more commercial HTAD panels. While this does not reflect the diagnostic yield of current HTAD panels due to the study’s ascertainment strategy, it highlights the limited overlap between current panel content and the broader spectrum of syndromic genes associated with aortic phenotypes.

One of the most prominent biological themes in genes with multiple cases of *de novo* PVs was the recurrence of chromatin-regulatory and epigenetic genes, which comprise a diverse group of disorders often classified as chromatinopathies, characterized by developmental delay, intellectual disability, congenital anomalies, and abnormal growth [23,24]. Although these disorders have historically been studied in the context of neurodevelopment, accumulating evidence suggests that vascular involvement may be underrecognized. For example, case studies of *NSD1*-associated Sotos syndrome have reported patients with aortic dilation, suggesting that aortic involvement may be more common than previously appreciated[9,25,26]. *DNMT3A*-associated Tatton-Brown-Rahman syndrome (TBRS) has also emerged as an important syndromic aortopathy, with recent studies documenting thoracic and arterial aneurysms, as well as aortic dissections and skeletal features of Marfan syndrome[10,27]. Our identification of additional rare *DNMT3A* variants in independent TAD cohorts, including the previously reported p.Arg635Gln variant in an individual with sporadic aortic dissection, provides further case-level support for an association between *DNMT3A* and aortic disease[10].

Aortic involvement was also observed across several chromatinopathies for which an association remains less well established. Aortic dilation has been reported in *EP300*-associated Rubinstein-Taybi syndrome (RTS). At the same time, our findings support an aortic phenotype in *CREBBP*-associated RTS, based on four unrelated GeneDx individuals with *de novo* PVs and four additional case-level observations in external TAD cohorts [28]. *ANKRD11* was the third most recurrent gene in the GeneDx cohort (*n* = 7), with emerging literature suggesting possible aortic involvement in KBG syndrome, although its extent remains uncertain [29,30]. *ADNP* was identified in five individuals in the GeneDx cohort, although one individual also harbored a *de novo KAT6A* PV. Independent support for *ADNP* was provided by a significantly increased burden of damaging LOF variants among TAA cases in AoU, despite the rarity of reported aortic involvement in Helsmoortel-Van der Aa syndrome[31]. Although TAD has not been reported in individuals with *KAT6A*- or *SRCAP*-associated disorders [32,33], additional rare, predicted-damaging variants in both genes were identified among individuals with TAD in the external, clinically ascertained cohorts. Aortic disease has likewise not been established as a feature of Coffin-Siris syndrome (CSS). However, we identified 14 individuals with aortic phenotypes across eight CSS-related genes (*ARID1A, ARID1B, ARID2, BICRA, DPF2, SMARCA4, SMARCB1*, and *SOX4*), most of which encode components or regulators of the BAF/SWI-SNF chromatin-remodeling complex[34,35]. Finally, an aortic dissection was recently reported in an adult with *KMT2B*-associated Kabuki syndrome[8]. Collectively, the recurrence of aortic phenotypes across multiple chromatin-regulatory disorders suggests that epigenetic dysregulation may contribute to aortopathy.

Genes in the RAS/MAPK signaling pathway harboring PVs were also identified in individuals with TAD phenotypes. Neurofibromatosis type 1 (*NF1*) is a well-established example, with reported thoracic and abdominal aortic aneurysms and aortic dissections[36]. *NF1* was supported by recurrent *de novo* variants in the GeneDx cohort, significant enrichment of damaging variants among dissection cases in the rare-variant burden meta-analysis, and an additional variant that segregated with TAD in an HTAD family. Aortic dilation has also been recognized in Noonan syndrome (*PTPN11*), with one cohort showing aortic root aneurysm in 21.6% of patients and progression over time[7]. Our primary *PTPN11* finding was supported by strong additional case-level evidence, with five cases identified across the HTAD, ESTAD, and trio cohorts carrying rare, damaging variants. Lastly, the *PPP1CB de novo* findings in the GeneDx cohort were supported by prior reports of aortic root dilation in Noonan-like syndrome and by significant enrichment of damaging missense variants among thoracic aortic aneurysm cases in the UKB [22].

Overgrowth syndrome genes were also identified with PVs associated with thoracic aortic disease codes. *NFIX*-associated Malan syndrome is characterized by overgrowth, macrocephaly, connective tissue manifestations, scoliosis, and pectus abnormalities, with reported cases of aortic root dilation and other cardiovascular findings[37]. *PIK3CA* and *PTEN*, both of which converge on the PI3K/AKT/mTOR pathway, are not established aortopathy genes[38]. However, *de novo* variants in both genes were identified in the GeneDx cohort, and *PIK3CA* had additional case-level support in HTAD pedigrees. Collectively, these findings suggest that dysregulated growth signaling may represent an emerging biological theme associated with an increased risk for TAD.

Several additional syndromic disorders not traditionally considered to cause aortic disease also emerged from the final analytic cohort. *ABL1 de novo* variants were identified in the GeneDx cohort, and germline *ABL1* variants have been associated with a syndrome characterized by congenital heart defects, connective tissue manifestations, skeletal abnormalities, and aortic root dilation in some affected individuals[39,40]. An individual with *PACS1*-associated Schuurs-Hoeijmakers syndrome and a sinus of Valsalva aneurysm has also been reported, and three unrelated individuals in the GeneDx cohort carried the same recurrent *PACS1* PV, suggesting this may be a common feature of this syndrome [41]. Lastly, *ZNF292 de novo* variants were also identified in the primary cohort, although this recently described neurodevelopmental disorder gene has no established association with aortic disease[42].

One of the most important observations from this study is the broad spectrum of syndromic genes associated with aortic phenotypes that are not represented on current commercial genetic testing HTAD panels. Many of these disorders demonstrate marked phenotypic variability, and characteristic developmental, craniofacial, behavioral, or overgrowth features may be subtle, age-dependent, or incompletely recognized, limiting the effectiveness of phenotype-directed testing strategies. Several also share clinical features, including tall stature, joint laxity, scoliosis, and connective tissue manifestations, that overlap with established aortopathy syndromes and may complicate clinical recognition. These findings support consideration of broader genomic approaches, including ES/GS, particularly in individuals with aortic disease accompanied by neurodevelopmental, syndromic, or multisystem features.

These data also have implications for cardiovascular surveillance. Thoracic aortic aneurysm was the predominant vascular phenotype in this study, whereas dissections were relatively uncommon. This difference may reflect the predominantly pediatric composition of the cohort. More than 80% of individuals were younger than 18 years at the time of testing. For TAD, aortic risk increases with age and aortic dilation precedes dissection [2,43]. At the same time, most genes identified in this study did not show a significant association with dissection in the biobank analyses, which contain primarily adults. It is notable that bicuspid aortic valve, a common congenital cardiovascular malformation, is associated with thoracic aortic aneurysm formation but confers a low risk of dissection[44]. Thus, a subset of the genes identified in this study, such as those involved in chromatin regulation, may affect developmental pathways that predispose to aortic aneurysm formation while conferring a low risk of aortic dissection. Nevertheless, longitudinal imaging studies are needed, and gene-specific natural history studies will be required to define penetrance, age-related risk, progression, and appropriate surveillance strategies.

There are several limitations of this study. Aortic phenotypes were identified through HPO-based ascertainment within a clinical laboratory database, and detailed imaging data were not uniformly available for review and/or validation of the reported phenotype. Consequently, the severity, progression, and anatomical distribution of vascular disease could not be systematically assessed. Ascertainment bias is also possible because individuals with neurodevelopmental and/or complex multisystem disorders may be more likely to undergo comprehensive clinical diagnostic testing by ES/GS. Additionally, because 45 cases were coded only as ‘aortic aneurysm’ without anatomic site, we could not determine whether they represented thoracic, abdominal, or mixed aneurysms. However, abdominal aortic aneurysm is predominantly a disease of older adults and is rare in children [45,46]. Therefore, it is unlikely that these pediatric cases were abdominal aortic aneurysms. Two individuals carried two PVs, raising the possibility of blended phenotypic effects, although the contribution of each variant cannot be determined from these cases alone. Lastly, while ES/GS broadens genetic evaluation beyond current HTAD panels, some clinically relevant genes, such as *PKD1*, remain technically challenging in standard workflows and may require specialized assays [47,48].

## Conclusions

This study demonstrates that aortic phenotypes occur across a diverse range of syndromic conditions extending well beyond the traditional spectrum of connective tissue disorders. Recurrent TAD-associated variants in genes involved in chromatin regulation, RAS/MAPK pathway, and overgrowth syndromes suggest that the genetic architecture of TAD is broader than currently appreciated. Among the genes evaluated, *DNMT3A, NF1, PTPN11, PPP1CB, NSD1, ANKRD11, ADNP,* and *ABL1* had the strongest evidence for aortic involvement, based on recurrent *de novo* PVs in the GeneDx cohort together with supporting biobank, external case-level, and/or published evidence. These genes warrant consideration for expanded aortopathy panels. Coffin-Siris/BAF-complex genes represent a distinct emerging group, with recurrent aortic phenotypes but more limited gene-specific evidence. These findings also support broader use of ES/GS, particularly in individuals with aortic disease and syndromic or multisystem features. Prospective studies are needed to define gene-specific penetrance, natural history, and surveillance recommendations.

## Data Availability

The data analyzed in this study were obtained from GeneDx under a collaborative data-sharing agreement. Because the data contain clinical genetic testing information and are subject to institutional and contractual restrictions, they are not publicly available. The UKB and AoU data are available to researchers upon approval by their respective access committees. The UTHealth datasets are available in dbGaP Study Accession: phs000693.v7.p3.

## List of Abbreviations

ACMG: American College of Medical Genetics and Genomics
AoU: All of Us Research Program
CSS: Coffin-Siris syndrome
ES: exome sequencing
FDR: false discovery rate
GS: genome sequencing
HPO: Human Phenotype Ontology
HTAD: heritable thoracic aortic disease
LOF: loss-of-function
MAF: minor allele frequency
MAPK: mitogen-activated protein kinase
OMIM: Online Mendelian Inheritance in Man
PV: pathogenic or likely pathogenic variant
RTS: Rubinstein-Taybi syndrome
TAA: thoracic aortic aneurysm
TAD: thoracic aortic disease
TBRS: Tatton-Brown-Rahman syndrome
UKB: UK Biobank.

## Declarations

### Ethics Approval and Consent to Participate

The research described in this manuscript conformed to the principles of the Helsinki. It was approved by the institutional review board (IRB) at the University of Texas Health Science Center at Houston (UTHealth Houston) and by GeneDx under an IRB-approved waiver of consent (WCG IRB, protocol 20162523), provided that only de-identified data from clinical testing were used.

### Consent for Publication

Written informed consent was obtained for participants in the UTHealth cohorts. The GeneDx cohort was analyzed under an IRB-approved waiver of consent using de-identified clinical data.

### Authors’ Contributions

D.R.M. and D.M.M. conceived the project. D.R.M., B.M., and D.G. analyzed the data. B.M. and K.M. contributed patient information. D.R.M., B.M., and D.M.M. wrote the manuscript and obtained input from other co-authors. All authors read and approved the final manuscript.

## Acknowledgements

We gratefully acknowledge the affected individuals who participated in this study, as well as the All of Us and UK Biobank participants, for their contributions, without whom this research would not have been possible. A large language model (ChatGPT, OpenAI) was used solely to assist with language editing and improving readability. RouteLLM (Abacus.AI) was used by GeneDx as a computational aid for organizing HPO terms into broad phenotype groups.

