## Supplemental Methods for "Thoracic Aortic Disease in Genetic Syndromes Beyond Established Aortopathy Genes"

**SUPPLEMENTAL MATERIAL**

**Supplemental Methods**

**Biobank Cohorts**

The UK Biobank (UKB) is a population-based study of approximately 500,000 individuals recruited across the United Kingdom [1]. The research presented here was conducted under UKB Application Number 75470 using data from release v18.1 (November 2023). We included genome sequencing data from participants of European ancestry, defined by self-report as “British,” “Irish,” “White,” or “Any other White background,” comprising approximately 95% of the UKB cohort.

The All of Us (AoU) Research Program is a large-scale initiative that collects health and genomic data from individuals across the United States [2]. For this study, we used the AoU v8 dataset, which includes genome sequencing data from >414,000 participants of diverse ancestral backgrounds. Complete biobank demographic data are provided in Table S5.

**Biobank TAD Phenotype Definitions**

TAD phenotypes were defined using diagnosis and procedure codes (Table S6), adapted from Klarin *et al.* [3]. Phenotypes comprised individuals with thoracic aortic aneurysm (TAA), aortic dissection (Dissection), thoracic aortic aneurysm requiring surgical repair (TAA-surgery), and a combined severe phenotype including all individuals with either dissection or TAA-surgery (Dissection + TAA-surgery). Controls were restricted to individuals without aortic disease and without ICD-10 codes for congenital malformations, deformations, or chromosomal abnormalities (Q00-Q99) to reduce potential confounding from syndromic conditions that may independently predispose to rare damaging variation. To ensure consistent phenotype definitions across cohorts, all ICD codes were harmonized to the UKB format, which omits decimal subcodes.

**Sequencing Data Processing, Quality Control, and Variant Annotation**

Sequencing data from the UKB, consisting of multi-sample project variant call format (pVCF) files generated with the Illumina DRAGEN pipeline (v3.7.8, aligned to GRCh38), were accessed via the UKB Research Analysis Platform and processed using custom Python scripts within that environment. Sequencing data from the AoU cohort, also processed with DRAGEN v3.7.8, were accessed via Hail within the AoU Researcher Workbench. Variant-level filters for both UKB and AoU included DRAGEN PASS status, QUAL ≥ 30, and <10% missingness. Genotype-level filters required read depth (DP) ≥ 10, genotype quality (GQ) ≥ 20, allele balance ≥ 0.2 for heterozygous genotypes, and exclusion of variants with excess heterozygosity.

Variant annotation and functional effect prediction were performed using Ensembl comprehensive protein-coding transcripts, and results were summarized based on MANE Select transcripts [4,5]. For the UKB cohort, we used bcftools, SnpEff, and SnpSift to annotate and extract qualifying variants from project VCF (pVCF) files [6–8]. Annotations included minor allele frequencies (MAF) from gnomAD v4 and pathogenicity predictions from dbNSFP v4.8 [9,10]. Identical filtering criteria were applied to the AoU cohort using pre-annotated Hail tables generated by Nirvana v3.18, with gnomAD MAF annotations provided through Nirvana and missense pathogenicity predictions from dbNSFP [11].

**Rare Variant Filtering Criteria**

Sequencing data were processed using standardized pipelines with rigorous quality control measures. For missense variant burden analyses, we applied stringent filters (gnomAD MAF < 8.16 × 10⁻⁶, REVEL > 0.649, and AlphaMissense > 0.2543) optimized for identifying variants of uncertain significance (VUSs) in HTAD genes associated with aortic dissections to identify rare, likely damaging variants across the 16 genes with three or more pathogenic or likely pathogenic variants in the primary GeneDx cohort [9,12–14]. Predicted LOF variant analyses were similarly restricted to rare variants (MAF < 8.16 × 10⁻⁶) with consequences including stop-gain, canonical splice-site, or frameshift indels in the 15 of the 16 genes (*ABL1, ADNP, ANKRD11, ARID1B, CREBBP, DPF2, KAT6A, NF1, NSD1, PACS1, PIK3CA, PPP1CB, PTPN11, SRCAP,* and *ZNF292*) with a Probability of Loss-of-function Intolerance (pLI) score ≥ 0.9 or ≤ 30 LOF alleles in gnomAD [9]. The combined burden of missense and predicted LOF variants was also evaluated, limited to the same LOF-intolerant genes.

**Rare Variant Burden Analysis**

Gene-based rare variant burden testing was performed separately in UKB and AoU using Firth’s penalized logistic regression, modeling case-control status as a function of rare variant carrier status with covariates for age, sex, and the first four (UKB) or five (AoU) genetic principal components [15]. Related individuals (kinship coefficient Φ > 0.1) and individuals with missing covariates or sex discordance were excluded, with cases preferentially retained. For autosomal genes, heterozygous and homozygous carriers were treated equivalently, and for X-linked genes, hemizygous males and heterozygous or homozygous females were considered carriers. For each gene, logistic regression was performed separately for each combination of variant class (missense, LOF, or combined missense + LOF) and TAD group (e.g., aortic dissection). Meta-analysis results from UKB and AoU were combined using an inverse-variance-weighted fixed-effects model to obtain an overall estimate under the assumption of a common genetic effect across cohorts [16].

For the genes observed in three or more individuals in the primary GeneDx cohort, multiple-testing correction was performed across all available biobank burden analyses for these genes. Raw P values from all tested combinations of cohort, aortic phenotype, and variant class were included in a single Benjamini-Hochberg false discovery rate (FDR) correction [17]. Analyses without a valid P value were excluded from the correction. FDR correction was performed before selection of the most significant result for each gene. For reporting, the analysis with the lowest raw P value for each gene was retained, along with its corresponding FDR-adjusted P value, where associations with FDR-adjusted P < 0.05 were considered significant.

**Additional Clinically Ascertained Cohorts**

Complete demographic data for these cohorts are provided in Table S5.

***UTHealth Heritable Thoracic Aortic Disease (HTAD) Cohort***
The UTHealth HTAD cohort included probands from 485 unrelated pedigrees, defined as families with ≥2 affected members with thoracic aortic aneurysm or dissection.

***Early-Onset Sporadic Thoracic Aortic Dissection and Additional Sporadic Trio Cohorts***
The Early-Onset Sporadic Thoracic Aortic Dissection (ESTAD) cohort included 564 individuals with sporadic thoracic aortic dissection. An additional 67 probands with sporadic aortic dissection underwent sequencing in a trio-based framework (proband, mother, father).

Exome or genome sequencing was performed on blood-derived DNA from affected individuals. For the case-level analysis, the 16 genes recurrently identified in the primary GeneDx cohort were evaluated for rare qualifying variants. All qualifying variants were required to have an MAF <5 × 10⁻⁵ in gnomAD v4.1.1. Missense variants were additionally required to have REVEL >0.5, whereas predicted LOF variants were evaluated in genes with pLI ≥0.9 and included stop-gain, canonical splice-site, and frameshift variants. In-frame insertions and deletions meeting the MAF criterion were also considered. The MAF threshold was selected to restrict analyses to ultra-rare variation compatible with dominant Mendelian disease, consistent with population-genetic estimates of maximum credible allele frequencies for rare Mendelian disorders and thresholds used in studies of inherited cardiovascular disease [18,19]. REVEL >0.5 was used as a permissive variant-prioritization threshold to retain sensitivity for potentially deleterious missense variants [12]. Because these cohorts were used to identify additional case-level observations rather than for formal gene-based burden testing, these broader criteria were applied as an initial screen, followed by manual review of phenotype, segregation, and variant-level evidence to determine whether a variant was retained as case-level supporting evidence.
